# Lower respiratory tract sampling via bronchoscopy in COVID-19 ARDS: A focus on microbiology, cellular morphology, cytology and management impact

**DOI:** 10.1101/2021.02.22.21252201

**Authors:** Sameer Bansal, Hariprasad Kalpakam, Ashwin Kumar, Anmol Thorbole, Amogha Varsha, Ravindra M Mehta

## Abstract

**Background:** Lower respiratory tract (LRT) sampling via bronchoscopy has been done sparingly in COVID-19 ARDS due to the high aerosol risk for the health care workers (HCW). Valuable information can be gained by detailed evaluation of bronchoscopic LRT samples.

**Methods:** LRT samples were obtained by bedside bronchoscopy performed in suspected or confirmed severe COVID-19 ARDS patients on mechanical ventilation. Only positive cases were included in the study. Microbiological, cellular and cytological studies including LRT COVID-19 RT-PCR were performed and analysed.

**Results:** 100 samples were collected from 63 patients, 53 were males (84%). 43 patients (68%) had at least 1 comorbidity. 55% cases had secondary bacterial infection as demonstrated by positive culture. Most of these infections were due to multi-drug resistant organisms (94.5%). The most common organisms were *Klebsiella pneumoniae* and *Acinetobacter baumanii* in 56.3% and 14.5% cases respectively. Fungal superinfection was observed in 9 patients (14.3%). Bronchoscopy helped confirm COVID-19 diagnosis in 1 patient and helped rule out COVID-19 in 3 patients who were eventually excluded from the study. The median BAL fluid (BALF) WBC count was 953 (IQR; 400-2717), with mean neutrophil count 85.2% (±13.9), and mean lymphocyte count 14.8% (±13.9). Repeat sampling done in some patients showed a progressive increase in the total WBC count in BALF, an increase in neutrophil percentage, and a higher chance of isolating an organism on the culture (81% repeat procedures were culture positive). The rate of super-infection increased with longer duration of illness. Patients with superinfection also had an increased WBC count (1001 vs 400), and lower lymphocyte percentage (19% vs 12% - OR - 6.8 [95%CI −14.3 − 0.7]). Bronchoscopic LRT sampling contributed significantly to modifying antibiotic coverage and discontinuing steroids in 37% cases.

**Conclusion:** This study describes a detailed analysis of bronchoscopic LRT sampling in critically ill COVID-19 patients. This provided important basic and applied information augmenting disease understanding and contributing to clinical management when there was scant information available in the pandemic.

## Introduction

Bronchoscopic lower respiratory tract (LRT) sampling in COVID-19 patients has been done sparingly due to the high aerosol-related risk to healthcare workers (HCW). (1,2) Very few studies have described microbiological, cellular & cytological aspects of LRT samples and their relationship with inflammation and infection in COVID-19 patients. This study analysed LRT samples obtained by bronchoscopy with a focus on the above aspects in mechanically ventilated (MV) critical COVID-ARDS (C-ARDS) patients.

## Methods

Retrospective study of 100 LRT samples collected by bronchoscopy performed for clinical indications at a tertiary committed COVID care center between August 25, 2020 and January 5, 2021, for MV C-ARDS patients with initially proven or later confirmed COVID-19.

Microbiological, cellular and cytological evaluation of these samples were done, and their management impact was analysed. Other relevant information recorded included demographic and clinical parameters including age, gender, duration of symptoms prior to hospitalization, presence of common co-morbidities and duration of ventilatory support prior to sampling.

## Statistics

Data was tabulated and analysed using SPSS (ver. 25.0, SPSS Inc). Results were analysed in a descriptive fashion as number and percentages, mean and standard deviation, median and inter quartile range (IQR). Difference between mean and medians was expressed using chi-square test and Mann-Whitney U-test respectively. Correlation analysis was done using linear correlation and results expressed using Pearson’s correlation coefficient. Statistical significance was taken at P <0.5.

## Results

100 bronchoscopic LRT samplings were done in 63 MV C-ARDS patients. 43 patients had one bronchoscopy procedure, while 20 patients had repeat procedures, for various indications such as clinical deterioration with new radiographic infiltrates, segmental collapse and hemoptysis.

### A. Microbiology

#### (i) Gram Stain

Gram’s stain showed pus cells in 79 cases (83%), with numerous pus cells (>25 per LPF) reported in 51/79 (64.5%) cases. Morphologically, copious purulent endobronchial secretions correlated with increased pus cells on BAL analysis.

Overall cellular analysis showed the median WBC count in BAL fluid as 953 (IQR; 400-2717). When there were many pus cells reported, median WBC count was 1628 - corresponding mean neutrophil count was higher with the presence of pus cells (89.2% vs 85.2%).

Gram stain could detect an organism in 36 cases (36%) - Gram negative bacilli (GNB) in 34 cases and Gram-positive cocci (GPC) in 2 cases, with culture positivity in 55 cases (55%).

#### (ii) Culture reports

Of the 100 cases, bacterial culture was positive in 55 cases (55%) with colony counts >105 CFU/ml and sterile in 45 cases (45%). All these patients were on prior antibiotics. *Klebsiella pneumoniae* & *Acinetobacter baumannii* were most commonly isolated organisms in 31 cases (56.3%) and in 8 cases (14.5%) respectively. Other organisms isolated were *Burkholderia cepacia* in 4 cases (7.2%), *Enterobacter cloacae* in 3 cases (5.4%), and *Acinetobacter iwoffii, Providencia stuartii and Serratia marcescens* in 2 cases each. *MRSA, Pseudomonas aeruginosa, Morganella morganii, Stenotrophomonas maltophilia and Citrobacter freundii* were other sporadically isolated organisms.

3 patients grew more than 1 organism in the BAL fluid. In 2 cases, these were *Klebsiella pneumonia*e with *Enterobacter cloacae*, while in 1 case it was *Acinetobacter baumannii* with *Burkholderia cepacia*. All but 3/55 cases had grown multi-drug resistant (MDR) organisms (94.5%), implying nosocomial superinfection.

#### (iii) Fungal evaluation

9 patients (14.3%) had fungal superinfection as determined by KOH mount, fungal cultures and/or BAL Galactomannan. 7 patients had a positive KOH mount, of which 5 showed budding yeast with septate hyphae, while 2 had presence of aseptate hyphae. Fungal culture was positive in only 2 patients, but majority of these patients were on empirical anti-fungal medications. One patient grew *Aspergillus niger*, and the other patient grew *Aspergillus fumigatus*. 4 of these patients with a positive KOH mount also had bacterial co-infection with MDR organisms, while 3 were bacterial culture sterile.

BAL galactomannan was sent for 6 patients and was elevated in all the cases. Galactomannan values were 1.65, 2.88, 1.72, 2.03, 1.52 and 2.14 in these 6 patients respectively (done by immune-enzymatic sandwich microplate assay; > 0.5 ODI considered positive). While 3 of these cases had a positive KOH mount, 3 did not stain with KOH. All 6 were culture negative. Appropriate anti-fungal agents were added in all the cases. Simultaneous blood cultures (±1 day) sent in these patients grew fungi in 3/9 patients - 1 patient grew *Candida auris*, and 2 *Candida albicans*.

#### (iv) COVID RT-PCR

In 1 patient, COVID-19 diagnosis was confirmed on BAL RT-PCR after two consecutive nasopharyngeal (NP) swabs were negative. Additionally, in 3 patients, COVID-19 was ruled out on LRT RT-PCR. An important observation was the duration of illness and persistence of RT-PCR positivity in the BAL of many patients. In 18/27 (67%) positive cases, BAL RT-PCR positivity duration was >10 days from the beginning of illness, in 14 cases (54%) it was >15 days, in 3 cases (11%) > 20 days, while 1 patient had a persistently positive BAL RT-PCR report for 67 days.

### B. Antibiotics

Sampling was done with prior broad-spectrum antibiotics on board, as per our clinical protocol for MV patients. Since majority of patients grew MDR organisms sensitive only to the polymyxin group of antibiotics, we changed our policy mid-way to empiric polymyxins for any patients suspected of new onset infection on MV. A new strategy also was the additional use of nebulized colistin via a closed, in-line nebulization circuit in patients who had copious purulent secretions in the airways. Subsequent culture reports confirmed MDR organisms in all these cases. After nebulized antibiotics, we found a reduction in the quantity and purulence of secretions in the majority of these patients.

### C. BAL cytology

The median WBC count was 953 (IQR; 400-2717), with mean neutrophils 85.2% (±13.9), and mean lymphocytes 14.8% (±13.9). 51% of patients showed ≥10% lymphocytes, 25.5% had ≥ 20%, 14.3% had ≥ 30%, while 6% patients had ≥ 40% lymphocytes in the fluid analysis. Mean neutrophil to lymphocyte ratio (NLR) in BAL was 13.3. The following findings were noted correlating duration of illness with BAL cytology:

1. As the duration of illness progressed, the mean lymphocyte count in BAL reduced, while the neutrophil count increased.
2. Pearson’s correlation coefficient between duration of illness and BAL lymphocyte count was - 0.27, an inverse relationship, though statistically insignificant. (p > 0.05)

We also analysed the peripheral blood sample drawn within 24 hours of the BAL. The mean peripheral WBC count was 17.4 (±7.2), with mean neutrophils 90.3% (±6.0) and lymphocytes 4.8% (± 4.5). Mean NLR ratio was 35. The correlation between BAL NLR and serum NLR was 0.1, a very weak positive correlation.

### D. Comparison of patients with and without super-infection

28/63 (44.5%) patients had a documented superinfection - bacterial, fungal or both. A comparative analysis of patients with and without superinfection is presented in Table 2. The average age of patients with superinfection was higher (64.3 vs 60.7 years), and superinfection was more common with chronic kidney disease (14.8% vs 8.8%), though statistically insignificant. All patients received steroids, 16/63 (25.4%) patients received tocilizumab, 8 in each group (29% vs 23%).

**Table 1:** Spectrum of infections on BAL.

| **Bacterial Infection Characteristics** |  |  |
| --- | --- | --- |
| Bacterial culture positive | 55/100 cases | 55% |
| MDR organisms | 52 | 94.5% |
| *Klebsiella pneumoniae* | 31 | 56.5% |
| *Acinetobacter baumannii* | 8 | 14.5% |
| *Burkholderia cepacia* | 4 | 7.2% |
| *Enterobacter cloacae* | 3 | 5.4% |
| *Acinetobacter iwoffii* | 2 | 3.6% |
| *Providencia stuartii* | 2 | 3.6% |
| *Serratia marcescens* | 2 | 3.6% |
| *Citrobacter freundii* | 1 | 1.8% |
| MRSA<sup>*</sup> | 1 | 1.8% |
| *Pseudomonas aeruginosa* | 1 | 1.8% |
| *Morganella morganii* | 1 | 1.8% |
| *Stenotrophomonas maltophilia* | 1 | 1.8% |
| **Fungal Infection Characteristics** |  |  |
| Fungal Infections | 9/100 cases | 9% |
| KOH mount positive | 7 | 77.7% |
| Galactomannan positive | 6 | 66.7% |
| Fungal culture positive | 2 | 22.2% |
| **COVID-19 RT-PCR** |  |  |
| BAL Positive | 27/38 cases | 71% |
| MRSA: Methicillin resistant Staphylococcus aureus |  |  |

**Table 2:**
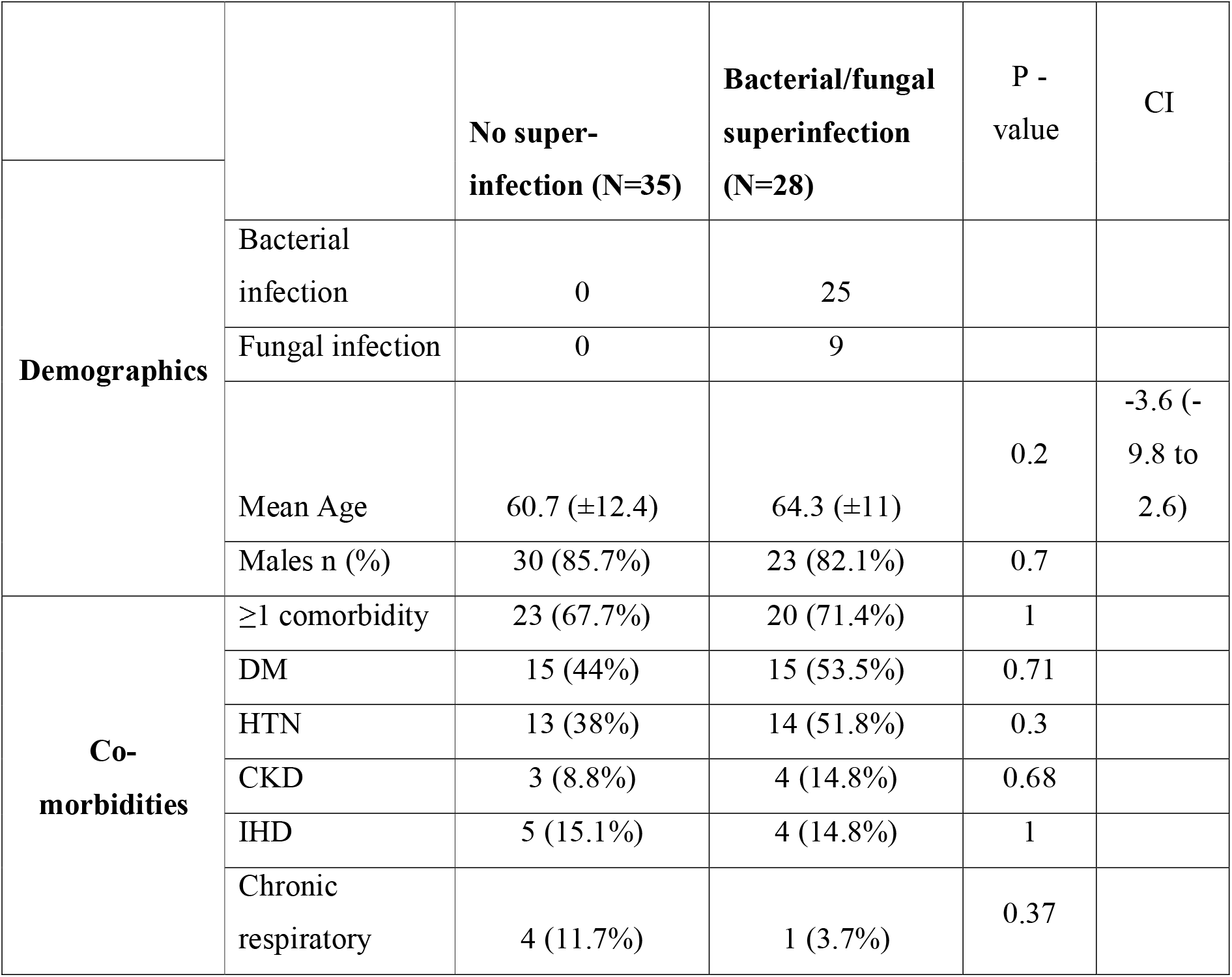
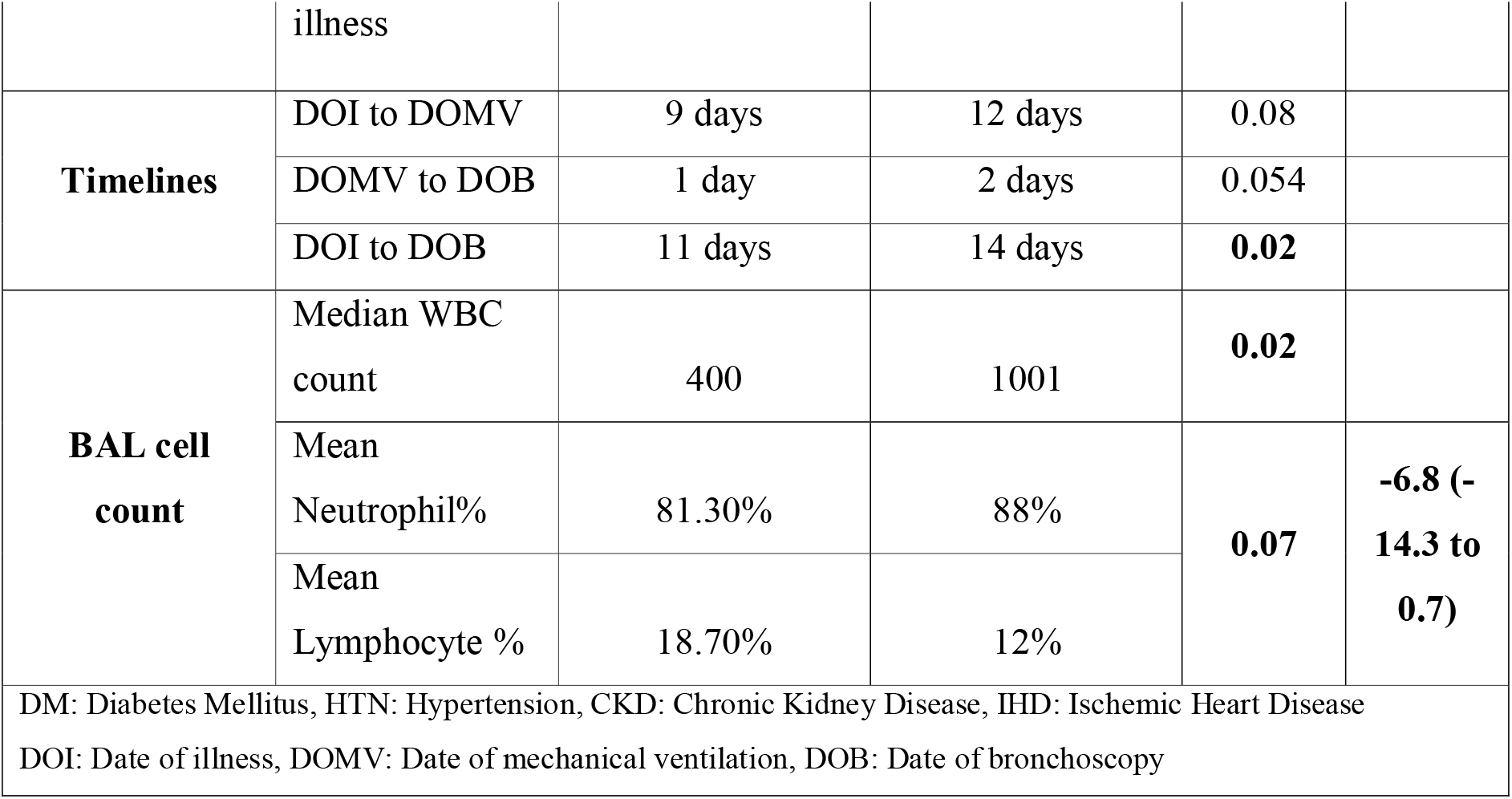
Comparison of patients with and without super-infection.

### E: Patients with repeat procedures

A subgroup of 20 patients had repeat bronchoscopy procedures with LRT sampling, majority for recurrent thick secretions in the endotracheal tube causing ventilation issues. The following aspects were noted. There was a greater chance of isolating an organism on culture when procedures were repeated (81% repeat procedures were culture positive). Serial changes noted on baseline broad-spectrum antibiotics included the following:

1. 14 patients had reduced secretion amount and purulence over serial procedures. WBC count also reduced sequentially in these patients. 4 patients turned culture negative on subsequent sampling, while 10 stayed persistently culture positive with the same microorganism with a significant colony count.
2. 6 patients had grown different microorganisms on serial bronchoscopic sampling. In these patients, WBC count in BAL fluid also increased sequentially, with the percentage of neutrophils increasing with repeat sampling.

### F. Multiple organisms at multiple sites

2 cases need special mention. One critical C-ARDS patient was put on extra-corporeal membrane oxygenation (ECMO) support. The second patient had a right bronchopleural fistula with a chest drain in situ. Both patients showed presence of multiple organisms at different sites simultaneously. The ECMO patient grew *Acinetobacter baumannii* and *Enterobacter cloacae* in the first BAL fluid with *Klebsiella pneumoniae* in the blood culture. Repeat LRT sampling after 4 days grew *Burkholderia cepacia*, while simultaneous blood culture grew *Pseudomonas aeruginosa*. The second patient had a BAL culture, blood culture and a pleural fluid culture. The BAL culture grew *Burkholderia cepacia*, pleural fluid grew *Pseudomonas aeruginosa* and the blood culture grew *Klebsiella pneumoniae*, all 3 within a short span. Both patients had been on MV for almost 15 days before being shifted to our facility and treated with prolonged high dose steroids (equivalent of 120 mg Solumedrol per day).

## Discussion

Limited data exists on detailed aspects of infection and inflammation in the lower respiratory tract in critically ill C-ARDS patients. This study of bronchoscopic LRT sampling in such patients describes noteworthy microbiological, cellular, cytological and RT-PCR aspects, and their clinical relevance in the pandemic. Additionally, the study reiterates the importance of detailed knowledge of local microbiological patterns in C-ARDS, vital to understand both disease dynamics and critical management issues such as superinfection.

LRT sampling revealed interesting aspects of bacterial super-infection. Reviewing published literature, Torrego et al found culture proven secondary bacterial infections in 28.6% cases (3), while cultures were positive in up to 60% cases sampled by Bruyneel et al (4). 86% of samples obtained by Baron et al. showed presence of at least 1 microorganism on culture (5). In our study, cultures were positive for various bacteria in 55% cases. The various microorganisms isolated reflects the local spectrum. In previous studies, the most common organisms isolated were *Pseudomonas spp*. and *Staphylococcus aureus*, while our series had majority positive for *Klebsiella pneumonia* and *Acinetobacter baumannii*. We also detected some uncommon pathogens viz. *Burkholderia, Providencia, Citrobacter, Morganella and Stenotrophomonas*, explained by advanced severe C-ARDS, comorbidities, and uniform steroid usage.

Fungal detection in LRT samples also varies depending on the series. COVID associated pulmonary aspergillosis (CAPA) has been described in multiple studies. Bruyneel et al reported fungal infection in 16 samples, all culture/galactomannan negative (4). Baron et al. isolated *Aspergillus spp*. on culture/PCR in 25% cases (5). Case series by Koehler et al. and van Arkel et al. suggest 20-25% incidence of Aspergillosis in critically ill COVID-19 patients (6,7). Studies from Wuhan reported secondary fungal infections in 35.3% critically ill patients (8,9). A case series from France reported presumed CAPA in 33.3% ICU COVID19 patients. (10) All-cause mortality was 33.3% in the French CAPA series and 80% in the study by Koehler et al. (10,6). Patrucco et al isolated fungi in 13% cases [*Candida albicans* 11 times (64.7%)]. (11) Previous studies have reported high rates of influenza-associated pulmonary aspergillosis (IAPA) which is similar to the high incidence of CAPA. (12) We had fungal superinfection in 9 (14.5%) patients. Since all our patients evaluated with BAL galactomannan had a value >1 with proven COVID, it seems reasonable to consider CAPA in these patients, even though they were culture negative.

Few studies describe the utility of LRT samples obtained by BAL for diagnosis of COVID-19 infection. This is an important clinical issue as the overall sensitivity of the nasopharyngeal (NP) COVID RT-PCR swab ranges from 55% to 70%. Wang *et al*. found SARS-CoV-2 RNA in 14/15 (93%) BAL samples in comparison to 126/398 (32%) of NP swabs from patients with COVID-19. (13) Patrucco et al. isolated SARS-CoV-2 27.5% times in patients with 2 negative swabs. (11) Similarly, LRT samples via bronchoscopy have also shown utility in ruling out COVID-19 disease. In a Roman study by Ora et al, BAL was done for confirmation of COVID-19 in patients with typical symptoms, suggestive CT scans, three consecutive NP/OP-negative swabs and IgG and IgM serology negative for SARS-CoV-2 (14). RT-PCR for SARS-CoV-2 was negative on LRT sampling in all 28 patients. The final diagnosis was pneumonia in 22, heart failure in 3, exacerbation of interstitial lung disease in 2 and ARDS in 1 patient. Antibiotic therapy was modified in 13 patients. We also found LRT sampling especially useful in this regard. BALF helped in diagnosing COVID-19 in 1 patient when 2 consecutive swabs were negative, while it helped in ruling out COVID-19 in 3 patients. These patients had a suggestive CT scan, elevated inflammatory markers, leukopenia, and 2 consecutive negative swabs. They were diagnosed as non-COVID viral pneumonia and eventually excluded from the study.

The SARS-CoV2 RT-PCR signal in the BAL and its relationship to symptom onset in critical C-ARDS is an important aspect that needs further exploration. Patrucco et al. performed BAL on 86 COVID suspects. Of these, 54 were RT-PCR negative and had a median symptom onset to bronchoscopy (SO→B) duration of 20 days. In comparison, 32 tested positive and had a median duration of SO→B of 12 days. (11) In our study, BAL RT-PCR was positive beyond 15 days in 54% of our cases, while the longest it remained positive was 67 days in one patient. Persistence of the BAL SARS-CoV2 signal has interesting implications on disease course, management and infectivity and needs further study, as it is difficult to assess whether the virus is dead or alive.

The cellular details in the LRT sampling showed interesting variations. The median WBC count in our study group was 953 (IQR; 400-2717), with mean neutrophils 85.2% (±13.9), and mean lymphocytes 14.8% (±13.9). As the duration of symptoms increased, lymphocyte percentage reduced, while neutrophils increased. This finding of BAL neutrophilia has been described in literature. Pandolfi et al. observed that alveolitis in severe COVID-19 patients was associated with hyperactivation of macrophages and neutrophils, with an excessive infiltration of neutrophils at the alveolar level (15). Lymphocytes were significantly reduced in critically ill patients as compared to patients admitted to the wards. Neutrophilia in these patients signifies severe inflammation with possible superinfection and portends worsening of disease with possible detrimental outcomes. Multiple autopsy reports have suggested role of neutrophilia as a marker of severe COVID-19. (16) All our patients were critically ill and had higher neutrophil percentage, with a mean NLR of 13.3. Barnes et al. proposed that neutrophilia could also be a source of excess neutrophil extracellular traps (NETs) - web-like structures of DNA and proteins expelled from the neutrophil that trap pathogens. (17) Liao et al. characterized BALF immune cells from patients with varying severity of COVID-19 and from healthy people by using single-cell RNA sequencing. (18) BALF of patients with critical COVID-19 infection showed a higher proportion of macrophages and neutrophils and lower lymphocyte count as compared to mild ones. Although most of the studies suggest high NLR as a poor prognostic model, none of them have commented on BALF NLR and its correlation with serum NLR. Mean serum and BALF NLR in our patients was 35 and 13.3 respectively. The correlation between BALF NLR and serum NLR was 0.1, a very weak positive correlation.

A unique aspect of our study was the analysis in the subset of patients with repeat procedures, average 5 days apart, for indications as mentioned above. Interesting findings in the LRT sampling in repeat procedures were a progressive increase in the total WBC count in BALF, decrease in the lymphocyte percentage, and higher chances of isolating an organism on the culture (81% repeat samples were culture positive).

Bronchoscopic LRT sampling results impacted management significantly. In 31.6% cases, antibiotics were escalated based on copious purulent bronchial secretions, with subsequent confirmation on culture. Analysis of preliminary culture results also led to a change in antibiotic policy with empirical polymyxin antibiotics introduced with suspicion of superinfection. Another unconventional strategy was the addition of nebulized colistin to systemic therapy. Other important decisions coinciding with antibiotic escalation were to deescalate or stop corticosteroids when copious purulent secretions were noted, as a systematic immunosuppression reduction strategy. Torrego et al. based on BAL introduced a new antibiotic in 83% patients. (3) Bruyneel et al. state that bronchoscopy led to antibiotic adaptation in 18% of total and 31% of positive microbiological samples. (4) Baron et al. mentioned that BAL impacted decision making in 71% cases: introduction, continuation, switch, or withdrawal of antimicrobial therapy in 50% cases, and decision to start (21%) or not start (21%) corticosteroid therapy. (5)

This is one of the few studies with comprehensive LRT sampling via bronchoscopy at the height of the COVID-19 pandemic, and correlates clinical, microbiological, cellular and RT-PCR findings. This is one of the few studies to report all these aspects with uniform steroid use. This data improves C-ARDS disease understanding, as well as help in clinical decision making in this critically ill population. Course corrections, such as altering the antibiotic strategy were implemented, and adding nebulized antibiotics was done as an additional strategy. The information gained from repeat procedures in some patients adds to the understanding of disease course. Our study did have certain limitations. We were restricted to only intubated critically ill COVID-19 ARDS patients, and not able to do galactomannan and molecular microbiological testing on all the samples.

## Conclusion

A fundamental limitation in MV COVID-19 patients was restricted suctioning due to aerosol risk, limiting many aspects of diagnosis and information to guide therapy. This study describes the detailed analysis and impact of bronchoscopic LRT sampling in critically ill C-ARDS patients at a stage when there was scant information available in the pandemic.

## Data Availability

All data is available for review

